# Critical Care for Severe COVID-19: A Population-based Study from a Province with Low Case-fatality Rate in China

**DOI:** 10.1101/2020.03.22.20041277

**Authors:** Xuelian Liao, Hong Chen, Bo Wang, Zhen Li, Zhongwei Zhang, Weimin Li, Zongan Liang, Jin Tang, Jian Wang, Rui Shi, Xiangde Zhen, Maojuan Wang, Xianying Lei, Yu Gong, Sheng Lv, Chao Jia, Li Chen, Juan Shang, Min Yang, Hailong Wei, Yuanjun Zhang, Xiong Yang, Huaqiang Shen, Xianhua Xiao, Jie Yang, Chang Liu, Qin Wu, Wen Wang, Jin Yang, Wanhong Yin, Xiaoqi Xie, Yongming Tian, Huan Liu, Bingxing Shuai, Wei Zhang, Xiangrong Song, Xiaodong Jin, Yan Kang, on behalf of the StUdy of 2019 Novel coRonavirus pneumonia Infected critically ill Patients in Sichuan provincE (SUNRISE) Investigators

## Abstract

**Background:** Data regarding critical care for patients with severe COVID-19 are limited. We aimed to describe the clinical course, multi-strategy management, and respiratory support usage for the severe COVID-19 at the provincial level.

**Methods:** Using data from Sichuan Provincial Department of Health and the multicentre cohort study, all microbiologically confirmed COVID-19 patients in Sichuan who met the national severe criteria were included and followed-up from the day of inclusion (D1), until discharge, death, or the end of the study.

**Findings:** Out of 539 COVID-19 patients, 81 severe cases (15.0%) were identified. The median (IQR) age was 50 (39-65) years, 37% were female, and 53.1% had chronic comorbidities. All severe cases were identified before requiring mechanical ventilation and treated in the intensive care units (ICUs), among whom 51 (63.0%) were treated in provisional ICUs and 77 patients (95.1%) were admitted by D1. On D1, 76 (93.8%) were administered by respiratory support, including 55 (67.9%) by conventional oxygen therapy (COT), 8 (9.9%) by high-flow nasal cannula (HFNC) and 13 (16.0%) by non-invasive ventilation (NIV). By D28, 53 (65.4%) were discharged, three (3.7%) were deceased, and 25 (30.9%) were still hospitalized. COT, administered to 95.1% of the patients, was the most commonly used respiratory support and met 62.7% of the respiratory support needed, followed by HFNC (19.3%), NIV ventilation (9.4%) and IV 8.5%.

**Interpretation:** The multi-strategy management for severe COVID-19 patients including early identification and timely critical care may contribute to the low case-fatailty. Preparation of sufficient conventional oxygen equipment should be prioritized.

**Trial registration number:** ChiCTR2000029758.

## INTRODUCTION

As a pandemic declared by the World Health Organization, the 2019 novel coronavirus disease (COVID-19) has affected over two million people worldwide since its outbreak by April 15, 2020 [1], in which approximately 19% are expected to progress to severe or critical disease, constituting the high risk group for death [2]. However, up to date, no population based data has been published regarding critical care for severe COVID-19.

The reported case-fatality rates among the severe COVID-19 varied a lot across different regions. Studies in Wuhan, including 13 to 199 severe cases identified using different criteria, reported high case-fatality rate from 19.2 to 61.5% [3–5]. Recent case series from Italy [6] that included 1591 cases reported a case-fatality rate of 26% while from US [7] that included 21 patients reported a rate up to 67%. Meanwhile, data from other regions outside Wuhan in China, reported much lower case-fatality rate ranging from zero to 6.5% [8–11]. In other countries with a relatively low case-fatality rate in all COVID-19 patients, very limited data were reported regarding severe cases. These studies provided valuable data regarding epidemiological, clinical, and biological characteristics, and treatment of the severe COVID-19. However, the reason for the discrepancy in fatality rate remained unclear. As no rigorous evidence has been published regarding the effectiveness of any treatment for COVID-19, it is likely that this discrepancy across regions could not be explained by the difference in the level of medical care or by any single therapeutic agent. Active management strategy and sufficient medical resource preparation have been proposed as two potential important factors for reducing the case-fatality rate of COVID-19 at the population level [12, 13]. However, real-world data are very limited regarding the clinical course, management, and medical resource usage for the severe COVID-19.

Sichuan province, located in the south west of China, covers a total area of 486 thousand square kilometres and is with 83 million population [14]. Since the first reported case of COVID-19 on January 16, 2020, a total of 539 cases have been confirmed in Sichuan and 3 deaths occurred among the severe cases, resulting in a 28-day case-fatality rate of 0.6% in all patients with COVID-19 and 3.7% in severe cases, which is much lower compared with that reported in most of the studies worldwide. Using data from the multicentre cohort study (StUdy of 2019 Novel coRonavirus pneumonia Infected critically ill patients in Sichuan provincE, SUNRISE), we aimed to describe and analyse the clinical course, multi-strategy management, and respiratory support resources usage for the severe COVID-19 at the provincial level.

## METHODS

### Study design and data collection

The SUNRISE study is a multicentre cohort study (http://www.chictr.org.cn/index.aspx, ChiCTR2000029758) focusing on COVID-19 in Sichuan Province, initiated by investigators in West China Hospital (WCH) [15]. All 21 hospitals designated for patients with severe COVID-19 in the province were included in the study. Data were prospectively collected for patients who were still in the hospital after study enrolment, and otherwise retrospectively collected, between January 16 and March 15. Daily data on COVID-19 cases reported in Sichuan province were obtained from Provincial Department of Health (SPDH) with permission. The timeline of the study is shown in the supplementary file. The study protocol was approved by the Ethics Committee of the West China Hospital and the participating hospitals. Informed consent was obtained from the patient or the patient’s legally authorized representative.

Detailed demographic, epidemiological, clinical and laboratory data were recorded using the electronic data capture and analysis system (EDC) (more details are provided in the supplementary file). Data entry was completed by physicians and nurses who were trained on the use of EDC and were working in the designated hospitals. Data quality was overseen by a team of senior ICU physicians and statisticians.

### Arrangement of designated hospitals and provisional ICUs

Considering the potential shortage of medical resources and region-related healthcare disparities, provisional ICUs were established to enlarge the capacity for receiving severe cases and ensure high quality care. 21 hospitals, one for each city, were planned as designated hospitals for severe COVID-19 patients in Sichuan province, after evaluating their capability of providing advanced respiratory support including high flow nasal cannula (HFNC), non-invasive (NIV), or invasive ventilation (IV). For the establishment of provisional ICUs, ICU physicians, nurses, respiratory therapists and resources were dispatched to designated hospitals under the organization of the authorities. The number of severe COVID-19 patients in each designated hospital was required to report daily. Remote multi-disciplinary consultation was arranged to discuss complicated cases daily.

### Definition of severe cases

COVID-19 was diagnosed using real time reverse transcription polymerase chain reaction (RT-PCR) of nasal, pharyngeal swab or sputum specimens by the local Centres for Disease Control (CDC). According to the criteria proposed by Chinese National Health Commission, confirmed COVID-19 patients who met any of the five following criteria [2] were included as severe cases: 1) dyspnoea or respiratory frequency ≥30 breaths/minute; 2) pulse oxygen saturation (SPO_2_) ≤93% without oxygen therapy in resting state; 3) PaO_2_:FiO_2_ ratio <300 mmHg; 4) lung infiltrates >50% within 24–48 hours; 5) respiratory failure, septic shock, and/or multiple organ dysfunction. Diagnosis of acute respiratory distress syndrome (ARDS) [16], acute kidney injury (AKI) [17], and septic shock [18] were made according to commonly used criteria (see supplementary file for details).

### Follow-up and outcomes

The day of enrolment of each patient with severe COVID-19 was considered day 1 (D1). Each patient included were followed up from D1 until discharge, death, or the end of the study. Patient characteristics were shown according to the outcome by D28 (see below). The main support methods used were analysed throughout the study period.

Clinical outcomes by D28, including rapid recovery (RR), prolonged recovery (PR) and no recovery (NR), were defined as follows. 1) RR: patient fully meeting the discharge criteria before D28, with normal body temperature ≥3 days, obvious improvement in respiratory symptoms and pulmonary imaging, and twice-negative nucleic acid tests (sampling interval being at least 24 hours) on respiratory samples; 2) PR: patient partially meeting the discharge criteria on D28 and still requiring hospitalization but without advanced respiratory support; 3) NR: death or the patient still in need of advanced respiratory support on D28.

### Statistics

Data management, manipulation, and analysis were conducted by a professional epidemiologist and statistician, from a third Clinical Research Centre, who did not participate in data collection. To ensure the high quality of database, all missing data and outliers detected were checked by two clinicians independently in the medical records. Then the verified data were collected and transferred for data completion or correction. No imputation was made for missing data. Data are expressed as median (IQR) for continuous variables and number (%) for categorical variables. For continuous variables, Wilcoxon test was applied to assess the difference between patients in one group and others, Kruskal-Wallis test to assess the overall difference among the RR, PR and NR groups. For categorical variables, χ2 test or Fisher’s exact test were performed according to the distribution of data. Two-sided tests with a significance level of 0.05 were applied. Needs for different respiratory support throughout the study period were assessed using person-day that denoted the use by one person in one day. All the analyses were conducted using R software version 3.6.2 (R Foundation for Statistical Computing). We used the tydyverse package [19] of to manage daily data and plot the daily use of main support and the pheatmap package [20] to plot the heatmap.

## RESULTS

### Early identification of severe COVID-19

Using the predefined criteria for severe COVID-19, 81 out of 539 patients were identified as severe cases. Among the remaining 458 patients with non-severe COVID-19, no death was observed during the study period. The median (IQR) durations from the onset of symptoms to the first hospitalization, RT-PCR confirmation and the diagnosis of severe condition were 3 (1–6), 7 (5-10), and 9 (6–11) days, respectively. Among the five criteria, PaO_2_:FiO_2_ ratio, SPO_2_, and dyspnoea criteria were the most commonly reported, accounting for 87.7%, 66.7% and 27.2%, respectively. On Day 1, all chest images of the patients (80 with CT scan and 1 with chest X-ray) showed bilateral lesions, though only 4 (4.9%) were diagnosed using the imaging criterion.

The main characteristics of the patients with severe COVID-19 were shown in Table 1. The median age (IQR) of the patients was 50 (39-65) years, 37.0% were female, and 50.6 % were with a body mass index (BMI) greater or equal than ≥24 kg/m^2^. Chronic comorbidities were observed among 43 (53.1%) patients. The Apache II [21] and SOFA score [22] were 10 (6-10) and 3 (3-5), respectively. 30 patients (37.7%) developed ARDS, six patients developed acute kidney injury (AKI) and one met the criteria for septic shock (**Table 1**).

**Table 1.** Clinical Characteristics of the severe COVID-19 patients, according to outcome by D28.

|  | **Total**<br>**(n=81)** | **Rapid Recovery**<br>**(n=53)** | **No Recovery**<br>**(n=10)** | **Prolonged Recovery**<br>**(n=18)** |
| --- | --- | --- | --- | --- |
| **Epidemiological and general characteristics** |  |  |  |  |
| Age, years | 50.0 (39.0-65.0) | 49.0 (39.0-63.0) | 76.0 (64.0-80.0) | 48.0 (37.0-60.0) |
| <45 | 28 (34.6%) | 20 (37.7%) | 6 (60.0%) | 8 (44.4%) |
| 45-65 | 30 (37.0%) | 21 (39.6%) | 3 (30.0%) | 6 (33.4%) |
| ≥65 | 23 (28.4%) | 12 (22.6%) | 1 (10.0%) | 4 (22.2%) |
| Sex | .. | .. | .. | .. |
| Women | 30 (37.0%) | 19 (35.9%) | 6 (60.0%) | 5 (27.8%) |
| Current smoking | 3 (3.7%) | 1 (1.9%) | 1 (10.0%) | 1 (5.6%) |
| BMI, kg/m<sup>2</sup> | 24.0 (21.5-27.3) | 24.5 (22.3-27.7) | 25.2 (21.1-27.3) | 23.2 (20.0-25.2) |
| <18.5 | 8 (9.9%) | 5 (9.4%) | 1 (10.0%) | 2 (11.1%) |
| 18.5-23.9 | 32 (39.5%) | 19 (35.9%) | 3 (30.0%) | 10 (55.5%) |
| 24-27.9 | 26 (32.1%) | 18 (34.0%) | 5 (50.0%) | 3 (16.7%) |
| ≥28.0 | 15 (18.5%) | 11 (20.8%) | 1 (10.0%) | 3 (16.7%) |
| Exposure to source of transmission | 62 (76.5%) | 42 (79.3%) | 6 (60.0%) | 14 (77.8%) |
| Family cluster | 19 (23.5%) | 15 (28.3%) | 1 (10.0%) | 3 (16.7%) |
| Living in or having travelled to Wuhan | 37 (45.7%) | 26 (49.1%) | 3 (30.0%) | 8 (44.4%) |
| Other scenarios <sup>b</sup> | 6 (7.4%) | 1 (1.9%) | 2 (20.0%) | 3 (16.7%) |
| **Symptoms** |  |  |  |  |
| Fever (≥37.5°C) | 39 (48.2%) | 25 (47.2%) | 5 (50.0%) | 9 (50.0%) |
| Non-productive cough | 40 (49.4%) | 29 (54.7%) | 3 (30.0%) | 8 (44.4%) |
| Productive cough | 38 (46.9%) | 24 (45.3%) | 3 (30.0%) | 11 (61.1%) |
| Fatigue | 30 (37.0%) | 20 (37.7%) | 2 (20.0%) | 8 (44.4%) |
| Dyspnea | 25 (30.9%) | 17 (32.1%) | 3 (30%) | 5 (27.8%) |
| Other symptoms <sup>c</sup> | 37 (45.7%) | 27 (50.9%) | 2 (20.0%) | 8 (44.4%) |
| **Coexisting disorders** |  |  |  |  |
| Any coexisting disorder | 43 (53.1%) | 23 (43.4%) | 10 (100.0%) | 10 (55.6%) |
| Diabetes | 18 (22.2%) | 8 (15.1%) | 3 (30.0%) | 7 (38.9%) |
| Hypertension | 15 (18.5%) | 9 (17.0%) | 4 (40.0%) | 2 (11.1%) |
| Chronic pulmonary disease | 11 (13.6%) | 5 (9.4%) | 3 (30.0%) | 3 (16.7%) |
| Cardiovascular or cerebrovascular disease | 5 (6.2%) | 2 (3.8%) | 3 (30.0%) | 0 (0.0%) |
| Congestive heart failure | 4 (4.9%) | 2 (3.8%) | 1 (10.0%) | 1 (5.6%) |
| Moderate to severe renal disease | 3 (3.7%) | 1 (1.9%) | 2 (20.0%) | 0 (0.0%) |
| AIDS, metastatic malignancy, or moderate to severe hepatic disease | 0 (0.0%) | 0 (0.0%) | 0 (0.0%) | 0 (0.0%) |
| Other coexisting disorder | 34 (42.0%) | 18 (34.0%) | 9 (90.0%) | 7 (38.9%) |
| **Clinical features on D1** |  |  |  |  |
| SOFA score | 3.0 (3.0-5.0) | 3.0 (3.0-4.0) | 7.0 (5.0-8.0) | 3.0 (2.0-4.0) |
| Apache II score | 10.0 (6.0-13.0) | 10.0 (6.0-12.0) | 15.0 (11.0-17.0) | 7.0 (2.0-10.0) |
| **Vital signs** |  |  |  |  |
| Systolic blood pressure, mmHg | 123.0 (114.0-137.0) | 123.0 (113.0-137.0) | 136.0 (121.0-158.0) | 121.5 (118.0-130.0) |
| Diastolic blood pressure, mmHg | 74.0 (68.0-80.0) | 75.0 (68.0-84.0) | 78.5 (73.0-80.0) | 71.5 (67.0-78.0) |
| Heart rate, beats/min | 88.0 (81.0-97.0) | 88.0 (81.0-97.0) | 87.0 (80.0-104.0) | 88.0 (83.0-95.0) |
| Respiratory rate, breaths/min | 22.0 (20.0-25.0) | 22.0 (20.0-25.0) | 22.0 (20.0-26.0) | 21.0 (20.0-24.0) |
| Pulse oxygen saturation, % | 96.0 (93.0-97.0) | 96.0 (93.0-97.0) | 96.0 (89.0-97.0) | 96.5 (95.0-97.0) |
| **Laboratory findings** |  |  |  |  |
| White blood cell count, × 10<sup>9</sup>/L | 6.2 (4.5-7.8) | 6.3 (4.9-8.2) | 6.7 (4.9-7.4) | 5.9 (3.9-9.5) |
| <4.0 | 17 (21.0%) | 10 (18.9%) | 2 (20.0%) | 5 (27.8%) |
| >10.0 | 11 (13.6%) | 7 (13.2%) | 0 (0.0%) | 4 (22.2%) |
| Neutrophil count, × 10<sup>9</sup>/L | 5.0 (3.1-6.6) | 4.6 (3.1-6.6) | 5.4 (4.5-5.7) | 4.2 (3.1-8.4) |
| Lymphocyte count, × 10<sup>9</sup>/L | 0.8 (0.6-1.1) | 0.8 (0.6-1.1) | 0.4 (0.3-1.1) | 0.8 (0.6-1.1) |
| <1.5 | 69 (85.2%) | 45 (84.9%) | 9 (90.0%) | 15 (83.3%) |
| Platelet count, × 10<sup>9</sup>/L | 166.0 (136.0-217.0) | 178.0 (139.0-237.0) | 124.0 (111.0-147.0) | 169.5 (150.0-202.0) |
| <150 | 31 (38.2%) | 19 (35.9%) | 8 (80.0%) | 4 (22.2%) |
| Total bilirubin, μmol/liter | 11.7 (6.9-18.4) | 12.6 (9.0-17.5) | 7.9 (3.5-19.5) | 10.8 (6.2-19.7) |
| Creatine, mg/liter | 67.8 (55.3-80.0) | 62.1 (53.8-78.9) | 80.0 (69.0-464.0) | 73.2 (59.9-84.0) |
| >133 | 5 (6.2%) | 2 (3.8%) | 3 (30.0%) | 0 (0.0%) |
| C-reactive protein, mg/liter <sup>c</sup> | 40.6 (18.7-69.3) | 39.4 (24.0-71.6) | 99.5 (43.0-129.0) | 33.0 9.92-44.6) |
| PaO<sub>2</sub>, mmHg | 69.2 (59.0-85.0) | 69.8 (58.8-84.3) | 71.6 (54.6-92.6) | 66.7 (60.0-104.1) |
| FiO<sub>2</sub>, mmHg | 33.0 (29.0-41.0) | 33.0 (29.0-41.0) | 33.0 (29.0-57.0) | 33.0 (29.0-37.0) |
| D-Dimer, mg/liter<sup>c</sup> | 0.8 (0.4-1.3) | 0.7 (0.4-1.2) | 1.3 (0.8-5.6) | 0.6 (0.2-1.1) |
| Bilateral lung infiltrates | 81 (100.0%) | 52 (98.1%) | 10 (100.0%) | 19 (100.0%) |
| **Criteria for the diagnosis of severe illness** |  |  |  |  |
| Dyspnea or respiratory frequency $\geq 30$ | 22 (27.2%) | 16 (30.2%) | 2 (20.0%) | 4 (22.2%) |
| Pulse oxygen saturation (SPO<sub>2</sub>%) $\leq 93\%$ | 54 (66.7%) | 34 (64.2%) | 6 (60.0%) | 14 (77.8%) |
| PaO<sub>2</sub>:FiO<sub>2</sub>, mmHg | 204.0 (156.8-266.5) | 204.0 (156.8-252.0) | 183.6 (115.8-255.2) | 224.8 (173.6-331.4) |
| <300 | 71 (87.7%) | 52 (98.1%) | 8 (80.0%) | 11 (61.1%) |
| <150 | 16 (19.8%) | 9 (17.0%) | 4 (40.0%) | 3 (16.7%) |
| Lung infiltrates >50% within 24 to 48 hours | 4 (4.9%) | 2 (3.8%) | 2 (20.0%) | 0 (0.0%) |
| Respiratory failure, septic shock, and/or MOF <sup>d</sup> | 17 (21.0%) | 7 (13.2%) | 5 (50.0%) | 5 (27.8%) |
| **Main organ dysfunction** |  |  |  |  |
| Acute respiratory distress syndrome | 30 (37.7%) | 20 (37.7%) | 4 (40.0%) | 6 (33.3%) |
| Acute kidney injury | 6 (7.4%) | 3 (5.7%) | 3 (30.0%) | 0 (0.0%) |
| Septic shock | 1 (1.2%) | 0 (0.0%) | 1 (10.0%) | 0 (0.0%) |
| **Main organ support** |  |  |  |  |
| No use | 5 (6.2%) | 3 (5.7%) | 1 (10.0%) | 1 (5.6%) |
| Conventional oxygen therapy through nasal catheter | 51 (63.0%) | 34 (64.2%) | 7 (70.0%) | 10 (55.6%) |
| Conventional oxygen therapy through mask | 4 (4.9%) | 3 (5.7%) | 0 (0.0%) | 1 (5.6%) |
| High flow nasal cannula | 8 (9.9%) | 5 (9.4%) | 0 (0.0%) | 3 (16.7%) |
| Non-invasive ventilation | 13 (16.0%) | 8 (15.1%) | 2 (20.0%) | 3 (16.7%) |
| Invasive ventilation or ECMO | 0 (0.0%) | 0 (0.0%) | 0 (0.0%) | 0 (0.0%) |
| Renal-replacement therapy | 5 (6.2%) | 0 (0.0%) | 5 (50.0%) | 0 (0.0%) |
| Vasopressors | 1 (1.2%) | 0 (0.0%) | 1 (10.0%) | 0 (0.0%) |

<sup>a</sup>. BMI was classified into 4 groups according to the criteria for Chinese population: underweight (<18.5 kg/m<sup>2</sup>), normal (18.5-23.9 kg/m<sup>2</sup>), overweight (24-27.9 kg/m<sup>2</sup>), and obese (≥28 kg/m<sup>2</sup>).
<sup>b</sup>. Other scenarios included having visited a designated hospital for COVID-19 patients or being in another city with confirmed COVID-19 cases during the past 14 days.
<sup>c</sup>. Other symptoms included muscle soreness, loss of appetite, diarrhea, abdominal pain, headache, dizziness, nausea, vomiting.
<sup>d</sup>. Data were missing for the measurement of C-reactive protein in 12 patients (14.8) and D-Dimer in four patients (4.9%).

### Centralization of patients and provisional ICUs for critical care

Out of the 21 planned designated hospitals for severe cases, 18 received severe patients during the study period. Among these hospitals, only 2 had standard ICU wards, while 16 had provisional ICUs which were transformed from general wards for infectious disease and equipped with ICU physicians and nurses. Under such situation, all 81 severe cases were centralized to the 18 designated hospitals, among whom 51 (63.0%) were treated in 16 provisional ICUs while 30 (37.0%) were treated in two standard ICUs. Only 11(13.6%) patients were admitted directly, 70 (86.4%) were transferred from non-designated hospitals or designated hospitals for non-severe cases. In total, 77 patients (95.1%) were admitted for critical care by D1 (Figure 1).

**Figure 1.**
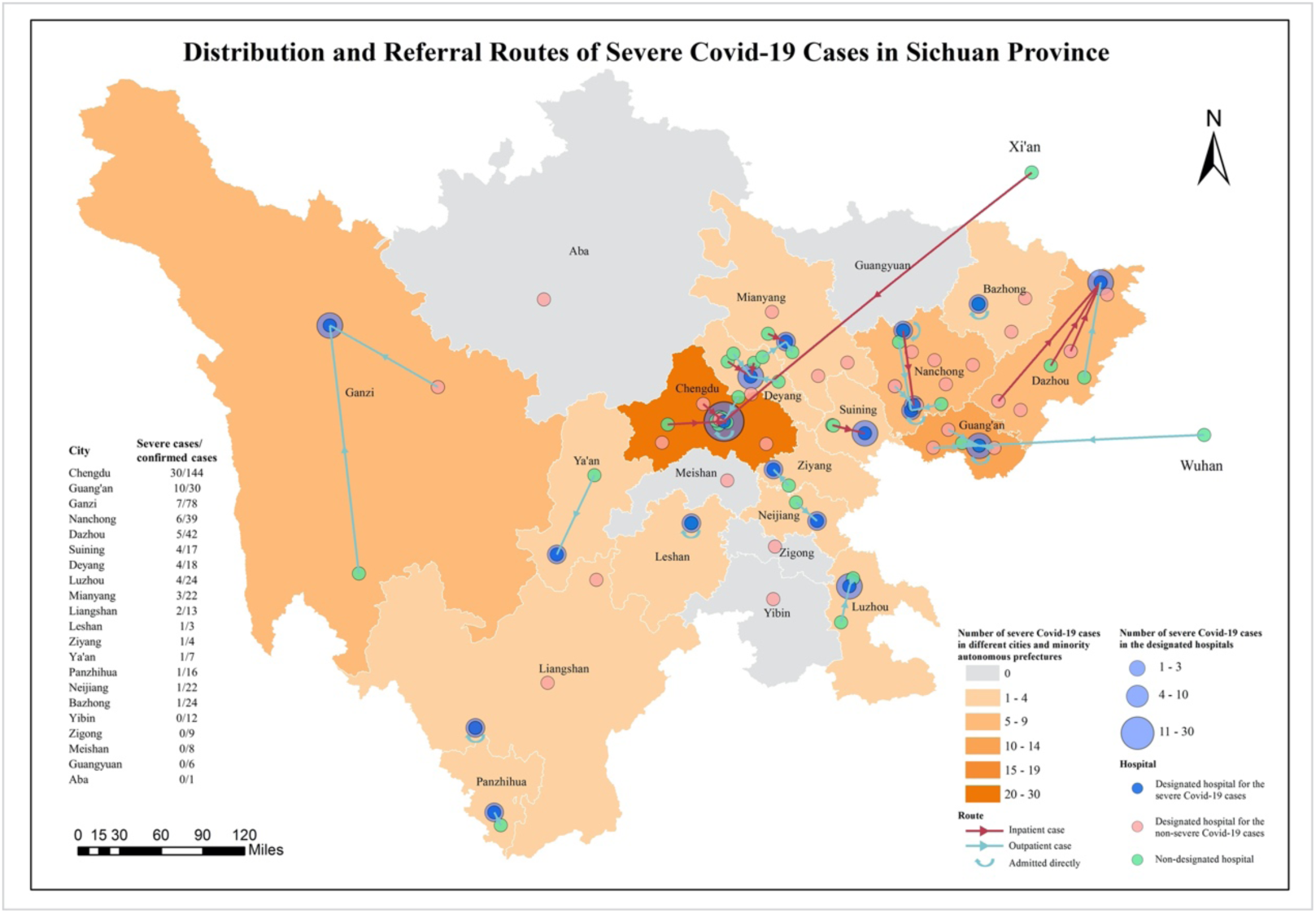
Distribution and referral routes of severe COVID-19 cases in Sichuan Province. Distribution of all the 81 patients with severe COVID-19 from 13 cities and 2 minority autonomous prefectures of Sichuan province were shown in the figure. The blue dots denote the 18 designated hospital for severe cases, pink dots denote 30 designated hospital for non-severe cases, and green dots denote non-designated hospital. The red and green arrows represent the route taken by inpatient and outpatient cases, respectively.

### Respiratory support during the study period

Respiratory support was the most commonly used organ support method for patients with severe COVID-19. On D1, 76 patients (93.8%) were administrated by respiratory support, including 55 (67.9%) by conventional oxygen therapy (COT) through nasal catheter or mask, 13 (16.1%) by NIV, and 8 (9.9%) by HFNC. No patient was intubated or needed ECMO (**Table 1**).

Daily respiratory support given to each patient from D1 to D28 were shown in **Figure 2**. Of the 81 patients diagnosed severe, 79 (97.5%) used COT, 31 (38.3%) used HFNC, 22 (27.2%) used NIV, 10 (12.3%) used IV, and 1 (1.2%) used ECMO (**Table 2**). Thirty-four patients (42% of 81) used only COT among which 79.4% discharged before D28. In the 25 patients who started with COT and needed escalation to advanced respiratory support methods, 12 (48.0%) were discharged by D28.

**Table 2.** Treatment for severe COVID-19 throughout the study period, according to outcome by D28.

|  | **Total**<br>**(n=81)** | **Rapid Recovery**<br>**(n=53)** | **No Recovery**<br>**(n=10)** | **Prolonged Recovery**<br>**(n=18)** |
| --- | --- | --- | --- | --- |
| **Respiratory Support** |  |  |  |  |
| Conventional oxygen therapy through nasal catheter | 78 (96.3%) | 52 (98.1%) | 8 (80.0%) | 18 (100.0%) |
| Conventional oxygen therapy through mask. | 10 (12.3%) | 6 (11.3%) | 2 (20.0%) | 2 (11.1%) |
| High flow nasal cannula | 31 (38.3%) | 18 (34.0%) | 6 (60.0%) | 7 (38.9%) |
| Non-invasive ventilation | 22 (27.2%) | 12 (24.5%) | 3 (30.0%) | 6 (33.3%) |
| Invasive ventilation | 10 (12.3%) | 0 (0.0%) | 7 (70.0%) | 3 (16.7%) |
| Extracorporeal membrane oxygenation | 1 (1.2%) | 0 (0.0%) | 1 (10.0%) | 0 (0.0%) |
| **Other support** |  |  |  |  |
| Parenteral nutrition | 17 (21.0%) | 14 (26.4%) | 3 (30.0%) | 0 (0.0%) |
| Prone position therapy | 16 (19.8%) | 11 (20.8%) | 3 (30.0%) | 2 (11.1%) |
| Duration of prone position therapy, day | 10.0 (4.0-19.5) | 10.0 (2.0-20.0) | 8.0 (5.0-19.0) | 17.0 (3.0-32.0) |
| Blood transfusion | 6 (7.4%) | 2 (3.8%) | 4 (40.0%) | 0 (0.0%) |
| **Drugs** |  |  |  |  |
| Lopinavir or Ritonavir | 71 (87.7%) | 47 (88.7%) | 6 (60.0%) | 18 (100.0%) |
| Abidol | 25 (30.9%) | 13 (24.5%) | 5 (50.0%) | 7 (38.9%) |
| Ribavirin | 12 (14.8%) | 9 (17.0%) | 0 (0.0%) | 3 (16.7%) |
| Nebulized recombinant human interferon $\alpha 2b$ | 57 (70.4%) | 32 (60.4%) | 9 (90.0%) | 16 (88.9%) |
| Thymosin $\alpha$ | 33 (40.7%) | 19 (35.8%) | 6 (60.0%) | 8 (44.4%) |
| Immunoglobulin | 15 (18.5%) | 10 (18.9%) | 2 (20.0%) | 3 (16.7%) |
| Methylprednisolone | 44 (54.3%) | 27 (50.9%) | 6 (60.0%) | 11 (61.1%) |
| Maximum dose, mg | 80.0 (40.0-80.0) | 80.0 (40.0-80.0) | 100.0 (80.0-160.0) | 80.0 (40.0-80.0) |
| Duration, day | 5.0 (4.0-7.0) | 5.0 (3.0-7.0) | 4.0 (3.0-8.0) | 6 (4.0-9.0) |
| Antibiotics | 58 (71.6%) | 39 (73.6%) | 8 (80.0%) | 11 (61.1%) |
| Analgesics or sedatives | 13 (16.1%) | 5 (9.4%) | 6 (60.0%) | 2 (11.1%) |
| Neuromuscular-blocking drug | 1 (1.2%) | 0 (0.0%) | 1 (10.0%) | 0 (0.0%) |

Data are median (IQR) or n (%).

**Figure 2.**
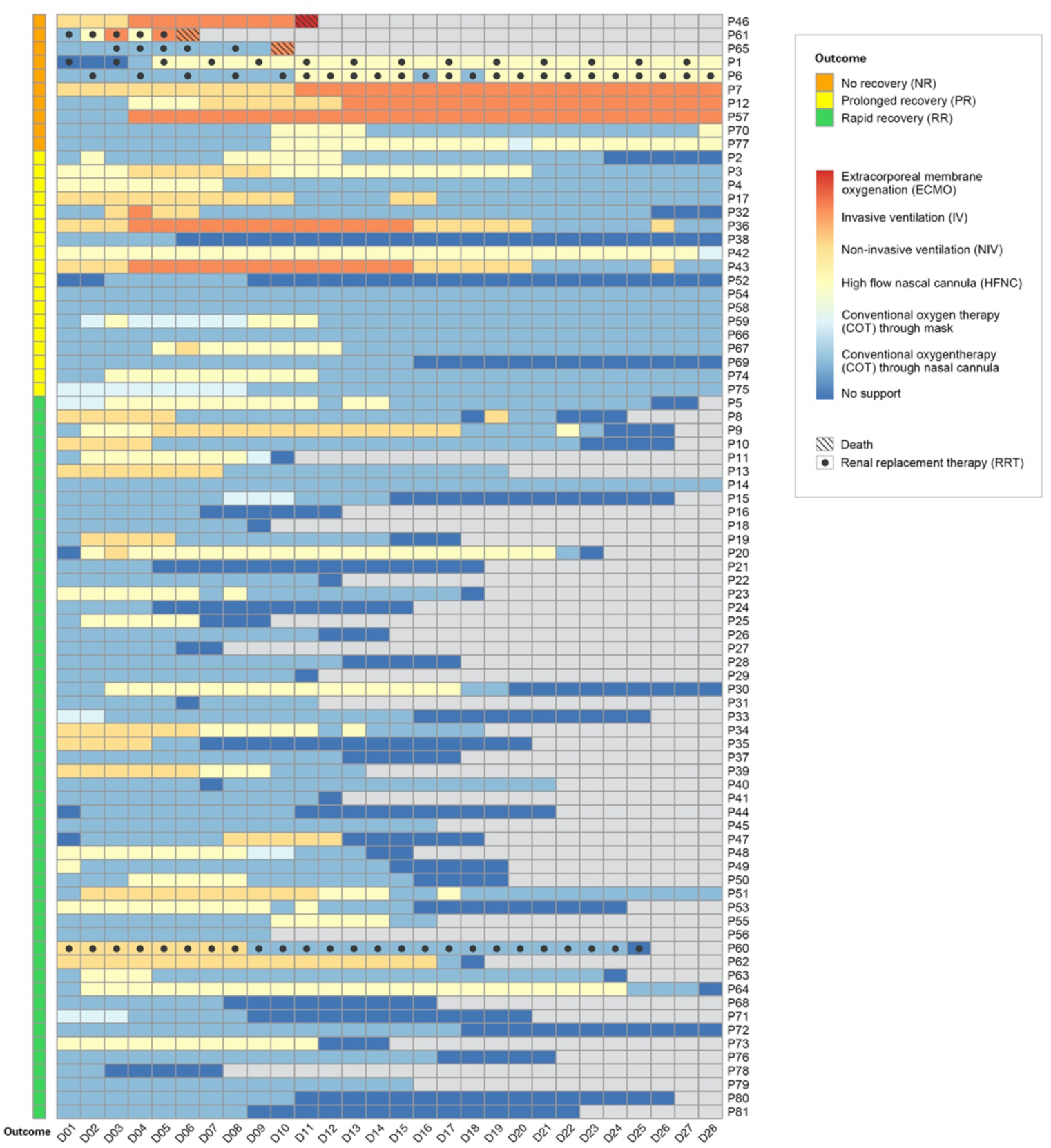
Daily organ support for 81 patients with severe COVID-19 from D1 to D28. Daily organ support, including respiratory support and renal replacement therapy, for each of the 81 patients with severe COVID-19 are shown from the day diagnosed severe (D1) to death, discharged from hospital, or D28.

Among patients who were discharged within 28 days, the median duration (IQR) of hospitalization was 18 (14–24) days. Only COT, HFNC, and NIV were used as respiratory support, for a median duration of 10 (5–14), 0 (0–4), and 0 (0–0) days, respectively. As for PR patients, COT, HFNC, NIV, and IV were used for 18 (7–24), 0 (0–7), 0 (0–3), and 0 (0–3) days, respectively. Concerning about the NR patients, the median duration for COT, HFNC, NIV and IV were 3 (1–10), 2.5 (0–11), 0 (0–3), and 2 (0–16) days, respectively. ECMO was used for one day by one patient.

We also analysed the usage of respiratory support along with the daily number of newly diagnosed patients with COVID-19 disease in Sichuan, newly diagnosed severe cases, and patients with severe disease hospitalized (**Figure 3**). In total, all forms of respiratory support were used 1579 person-day, of which COT took up 62.7 % (990 person-day), HFNC 19.3% (305 person-day), NIV 9.4% (149 person-day), IV 8.5% (134 person-day), and ECMO 0.06% (1 person-day). The peak needs of respiratory support, which had a significant lag of 9 days behind the peak of newly diagnosed patients in Sichuan, lasted for 20 days and paralleled with hospitalization needs for severely ill patients. During the most demanding days for respiratory support measures, 55.3% demands were COT, 21.3% were HFNC, 12.8% were NIV, and 10.6% were IV.

**Figure 3.**
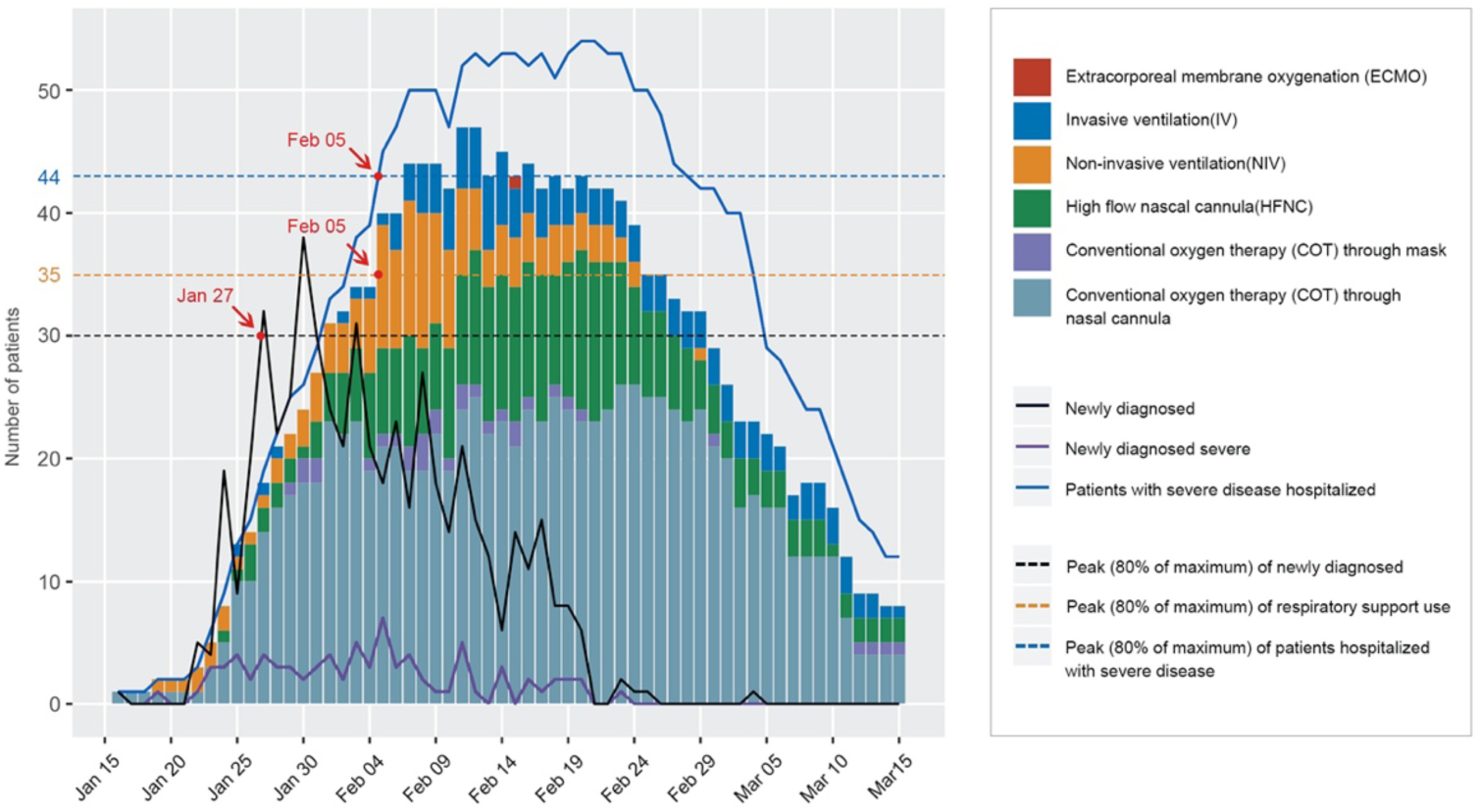
Daily respiratory support needs for patients with severe COVID-19 from January 16 to March 15. The bar plot shows, for each calendar day, counts of the respiratory support used for patients with severe **COVID**-19. Daily number of newly diagnosed patients with COVID-19 disease in Sichuan, newly diagnosed severe cases, and cumulative severe cases hospitalized are shown in lines.

### Other support measures and medical treatment

Some patients needed life-saving measures such as renal replacement (five patients, 6.2%), vasopressors (5, 6.2%), and blood transfusion (6, 7.4%). Other support methods included the prone position, partial parenteral nutrition therapy, analgesics and sedatives, used by 16 (19.8%), 17 (21.0%), and 13 (16.1%) patients, respectively. Various drug treatments were given despite the absence of effective evidence-based antiviral and immunomodulates (**Table 2**).

### Patient follow-up and clinical outcomes

All patients were followed up to the end of the study. Among 81 patients, 53 (65.4%) were discharged before D28 and regarded as rapid recovery (RR). Eighteen (22.2%) patients were regarded as prolonged recovery (PR), including 13 still in need of conventional oxygen therapy and 5 awaiting negative results of RT-PCR on D28. Ten patients (12.3%) were in the no recovery (NR) group, including 3 deaths and 7 still in need of advanced respiratory support on D28. Among the 3 deceased, one was a 64- year-old female with scleroderma, pulmonary fibrosis and diabetes, one was an 80- year-old female with hypertension and coronary heart disease. Both patients developed severe respiratory failure and died of multi-organ dysfunction. The third patient was a 73-year-old male with hypertension and end-stage renal disease and died from circulatory failure.

Characteristic of the patients on D1 according to the outcome were reviewed in **Table 1**. Patients in the NR group tended to be older (p<0.001), had higher Apache II (p=0.01) and SOFA scores (p<0.001), and were more likely to have comorbidities (p=0.02) including AKI (p=0.004) than the other groups. C-reaction protein (p=0.02) was higher while lymphocyte (p=0.03) and platelet (p=0.005) were lower in the NR group (**Table 1**).

By March 15, among those who were still hospitalized on D28, 15 were discharged, 9 patients were still hospitalized, and an 81-year-old male died of end-stage chronic pulmonary disease on D45.

## DISCUSSION

To our knowledge, it is the first time that critical care for severe COVID-19 was described from the perspective of identification, organization, and resources using data from a population based study, where a significant lower case-fatality rate was observed than that reported elsewhere [3–5, 7, 23]. Using the five predefined criteria, severe cases were identified early, none of cases needed mechanical ventilation or ECMO on D1. Providing timely critical care by establishing provisional ICUs, conventional oxygen therapy was the most commonly used respiratory support method given to 95.1% of the patients and met 62.7% of all the respiratory support needed.

Early identification of severe COVID-2019 is a prerequisite for timely interventions for severe COVID-2019. For the diagnosis of severe cases, different criteria were used in the previous studies while some didn’t reported clear criteria [3–5, 7, 23–26]. In this study, using the severe criteria proposed by Chinese National Health Commission [2], none of the non-severe patients died during the study period, and none of severe cases needed mechanical ventilation or ECMO on the day of diagnosis. In particular, our data showed that, on the day of diagnosis of severe, up to 72.8% did not present symptoms of dyspnoea and 33.3% had an SPO_2_ no less than 93%. Using only dyspnoea or SPO_2_ as the diagnostic criteria would fail to identify a large number of severe cases of COVID-19 in early stage and miss the opportunity of intervening in time. With a sensitivity of 87.7%, PaO_2_:FiO_2_ ratio was likely a sensitive indicator for early identification of severe COVID-2019. With early identification of the severe illness, 93.8% patients in this study were receiving various respiratory support by D1, with a median PaO_2_:FO_2_ ratio of 204 mmHg. In the previous studies, the reported median PaO_2_:FO_2_ ratio on ICU admission was much lower (103.8 to 169.0 mmHg) [5, 7, 27], while many other studies didn’t report the data. Similar to the strategy applied in Sichuan, all the patients outside Wuhan were less severe as demonstrated in Beijing, Shanghai, Zhejiang, Jiangsu, and Shanxi in China [28]. The use of sensitive diagnostic criteria might be crucial for improving prognosis for patients with old age or comorbidity, for whom symptoms may be more atypical and the progression of the disease may be faster than others.

Shortage of beds is one of the most challenging issues in fighting severe COVID-19 [29]. Despite very limited evidence reported in the literature, the data in our study showed that provisional ICUs may be a practical solution. By the establishment of the provisional ICUs, the critical care capacity for COVID-19 had a significant increase at the provincial level. The beds, organ support equipment, and staffs were arranged by the number of patients and adjusted over time. Provisional ICUs were not comparable to standard ICUs, but basic critical care, including main organ supports, could meet the majority needs of patients with severe COVID-19 and be provided in time. As a result, all the patients in this study received critical care in provisional ICUs, while in another study, only 19.1% of the severe cases could be treated in ICUs [23]. In regions without stand ICUs are not available or with a shortage of resources due to outbreak of the disease, provisional ICUs may alleviate the pressure of patient flow and provide timely critical care for patients with severe COVID-19.

The shortage of advanced support equipment such as ventilators is another challenging issue [30]. As there’s no definite effective drug to treat COVID-19, appropriate respiratory therapy is essential for severe cases. According to data reported in studies from Wuhan and a study from the US, IV was administered in 38.9-71% patients [7, 23, 27]. However, in our study, IV was used in only 12.3% of the patients and the case-fatality rate was much lower than that in these studies. This difference may be explained, at least in part, by the timing of intervention [12]. It is reasonable to hypothesize that hypoxemia may participate in multiple organ injury if the hypoxic compensatory period was overlooked or not treated timely [31]. Our study showed that, if severely ill patients can be identified earlier, traditional oxygen therapy and close monitoring may suffice, as in the 42% of the patients in our study. Given that the COVID-19 outbreak is moving rapidly, this finding is of particular interest for treating newly diagnosed severe cases and for regions facing a shortage of ventilators. Preparing a large availability of conventional oxygen therapy and providing it in an early stage, in hospital or even at home, might be the most cost-efficient alternative for improving the prognosis of severe COVID-19 patients and to relieve the burden of the whole medical system.

Our study has the following limitations. First, a small part of the data was collected retrospectively, leading to potential incompleteness and inaccuracy for some variables. We mitigated this limitation by designating a team of researchers to verify and complete the data. Second, our study was mainly descriptive due to the small sample size, and more rigorous evidence is needed for illustrating the benefits of using COT in early stage. Third, the findings reported in our study might not be generalizable to populations with completely different impact of the pandemic and government strategies.

Multi-strategy management, including early identification and timely critical care provided by establishing provisional ICUs is feasible and effective in treating severe COVID-19. Respiratory support, conventional oxygen therapy in particular, should be sufficiently prepared.

## Data Availability

Not Applicable.

## AUTHOR CONTRIBUTIONS

All authors contributed to the study design, conduction, data collection, writing, examination and approval of the final manuscript.

Yan Kang, Weimin Li, Zongan Liang, Xuelian Liao, Hong Chen, Bo Wang, Xiaodong Jin, Zhen Li, Weizhong Zhang, Qin Wu, Jin Yang, Chang Liu, Wanhong Yin, Xiaoqi Xie, Yongmin Tian, Wei Zhang, Huan Liu, Xiangrong Song, Bingxing Shuai, and Wen Wang conceived and designed the study; Yan Kang, Xuelian Liao, Qin Wu, Jin Yang, Bo Wang, and Wanhong Yin wrote the protocol for project of novel coronavirus pneumonia in West China Hospital; Xiaodong Jin, Weizhong Zhang and Zongan Liang contacted with each centre and acquired the data from the government; Hong Chen, Jin Tang, Jian Wang, Rui Shi, Xiangde Zhen, Maojuan Wang, Xianying Lei, Yu Gong, Chao Jia, Sheng Lv, Li Chen, Juan Shang, Min Yang, Hailong Wei, Yuanjun Zhang, Xiong Yang, Huaqiang Shen, and Xianhua Xiao were responsible for the local research progress and ensure the data quality; Xuelian Liao, Jie Yang, and Wanhong Yin contributed to database creation and standardization; Zhen Li, Xuelian Liao, and Jie Yang analysed data or performed statistical analysis; Yan Kang, Xuelian Liao, Zhen Li, Bo Wang, and Xiaodong Jin wrote the manuscript and had primary responsibility for final content.

## ACKNOWLEDGEMENT

We would like to thank Ms. Haixin Miao, Mr. Desong Qiu from Sichuan Zhikang Technology CO., Chengdu, for their help of establishing the electronic data capture and analysis system; We thank Ms. Yu Ma, Mr. Biwei Zhan from Chengdu Urban Planning Information Technic Centre for the localization and mapping for the designated hospital. We also thank Yi Liu, Ph D. from Shanghai, for his assistance in data visualization.

